# Relationship between psychosocial factors and cardiovascular autonomic function among junior doctors: A cross-sectional study

**DOI:** 10.1101/2025.10.12.25337844

**Authors:** Kofi Tekyi Asamoah, Richard Dei-Asamoa, Eugene Baafi Ampofo, Felix Razak Awindaogo, Francis Agyekum, Alfred Doku, Sammy Ohene, Eugene Amable

## Abstract

**Background:** Psychosocial factors such as stress, anxiety, and depression, are important causes of CVD by contributing to autonomic dysfunction, predisposing to endothelial dysfunction, arrhythmogenesis, among others. Doctors are at a high risk of psychosocial distress due to factors such as heavy workload and taking end-of-life care decisions. We sought to determine the prevalence of psychosocial factors among junior doctors and explore their relationship with cardiovascular autonomic function.

**Methods:** A cross-sectional study was conducted among junior doctors at the Korle-bu Teaching Hospital from April to August 2023. Data were collected using an electronic questionnaire and a treadmill stress test. The Depression, Anxiety, and Stress Scale −21 items (DASS-21) was incorporated into the questionnaire to assess depressive, anxiety and stress symptoms. Cardiovascular autonomic function was determined by heart rate recovery and chronotropic index. The data collected were analysed with the Stata version 16.1, using descriptive and inferential statistics.

**Results:** 244 junior doctors participated in the study, with a mean age of 31.46. The prevalences of depressive, anxiety and stress symptoms were 29.51%, 48.77% and 48.36% respectively, correlating moderately with one another. Chronotropic index was abnormal for 9.43% of respondents (95% CI = 6.07% - 13.81%) and heart rate recovery-at-1-minute (HRR_1_), 3.28% (1.43% - 6.36%). There were no differences in autonomic function profile based on tertiles of psychosocial factors using the Kruskal-Wallis test and one-way analysis of variance. Stress scores were associated with increased HRR_1_ (β per SD 1.53 bpm, 95% CI 0.03 - 3.02). Higher depression scores were associated with lower odds of chronotropic incompetence (adjusted OR 0.44, 95% CI 0.22 - 0.88).

**Conclusion:** There is a high prevalence of depressive, anxiety and stress symptoms among junior doctors. This is however associated with augmented rather than impaired cardiovascular autonomic function.

## Introduction

Cardiovascular disease (CVD) is a leading cause of morbidity and death in sub-Saharan Africa (SSA) [1], accounting for a significant disability-adjusted life year (DALY) burden [2]. CVD rates have increased in low- and middle-income countries (LMICs) by up to 50% over the last 3 decades [1,2].

Psychosocial stressors, which include stress, anxiety and depression, contribute to CVD symptoms acutely through sympathetic nervous system (SNS) activation [3,4], and chronically through various mechanisms [4] (Fig. 1). They also worsen pre-existing CVD like heart failure and myocardial infarction [5]. Acute stressors evoke a physiological response through the SNS [4,6], followed over time by a psychological response [7]. The physiological response to stress involves two major biochemical pathways; the SNS and the hypothalamic-pituitary-adrenal (HPA) axis [4,6–8]. If personal coping mechanisms are unable to resolve the psychosocial disturbance, the chronic SNS activation in response to stress will result in noradrenaline secretion, activation of the renin-angiotensin-aldosterone system (RAAS), and the elaboration of inflammatory cells and cytokines [8,9]. Conversely, the parasympathetic nervous system (PSNS) output diminishes [7], upsetting sympathovagal balance and increasing resting heart rate (HR) and blood pressure (BP) [4,10]. The resting HR may therefore be an index of one’s cardiovascular autonomic function, though Lankhorst et al.[11] and Bustamante-Sánchez et al.[12] identified heart rate recovery (HRR) as an earlier and more sensitive sign of autonomic disturbance than resting HR (Fig 1).

**Fig 1.**
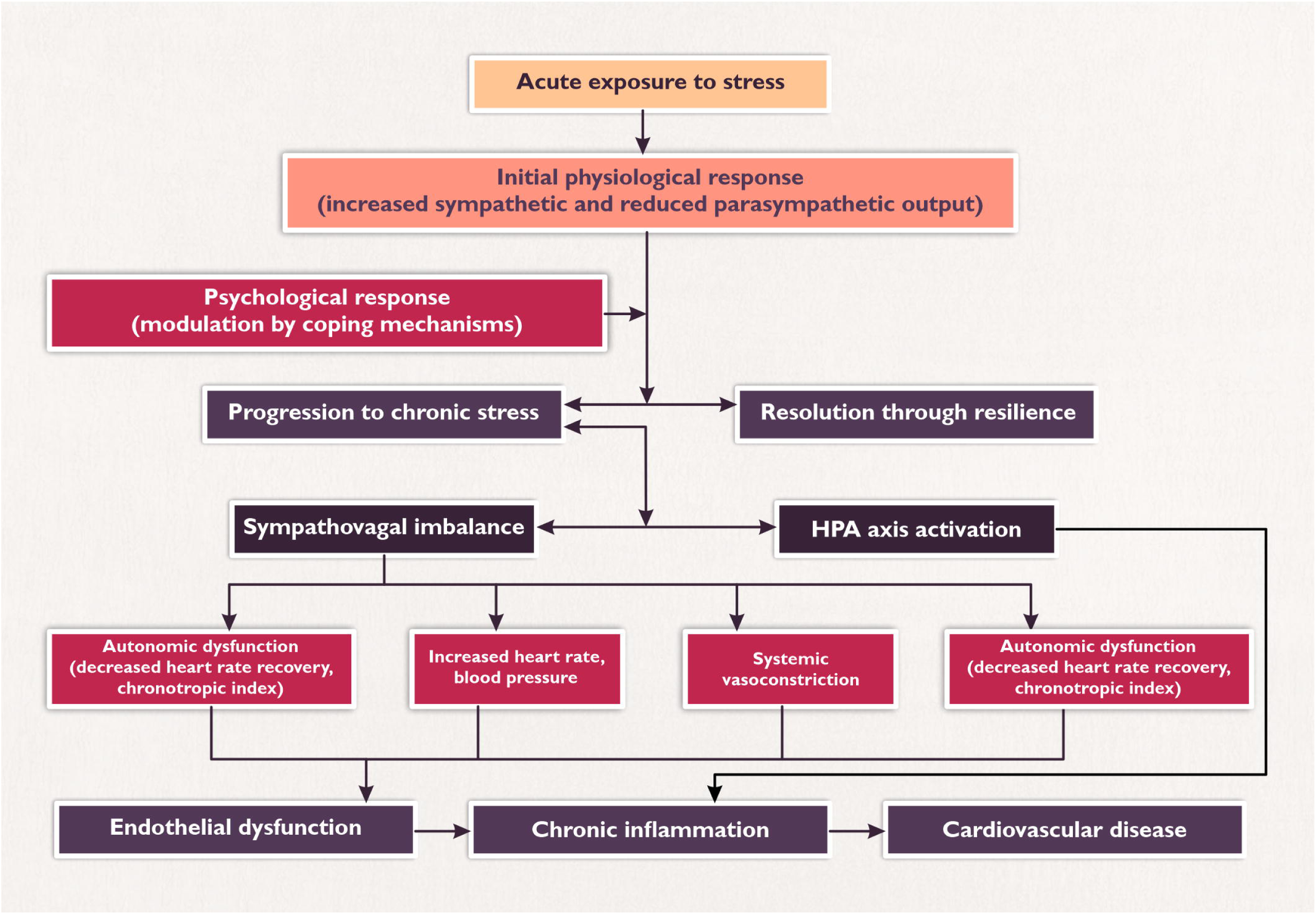
Diagram showing the study’s conceptual framework from psychosocial stress exposure to development of cardiovascular disease

**Fig 2.**
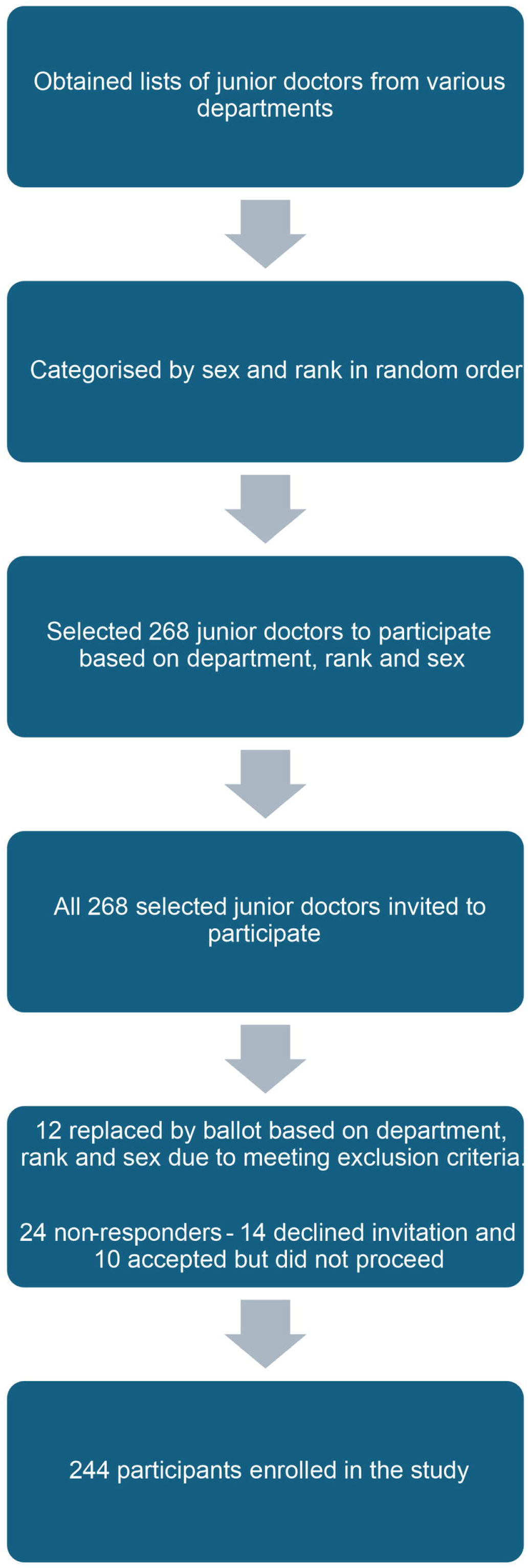
Flow chart showing the recruitment process for the study

SNS-induced vasoconstriction alters laminar vascular blood flow [6] through reduced endothelial nitric oxide synthase (eNOS) activity [13]. Cortisol, produced by the HPA axis, also inhibits eNOS [13]. Nitric oxide depletion favours platelet aggregation, elaboration of E-selectin and vascular cell adhesion molecule [14], creating a proinflammatory milieu that leads to endothelial dysfunction and increased atherogenesis [4,14]. The INTERHEART study reported that psychosocial factors confer an almost 3-fold increase in risk of acute myocardial infarction [15]. The INTERSTROKE study also reported that permanent or repeated episodes of stress increased the incidence of stroke, more with occupational stress than other forms [16].

Sympathovagal imbalance may be assessed by various non-invasive means. These include HRR and chronotropic index [17]. HRR is assessed using the treadmill stress test (TMT) and is the difference between HR at peak exercise and HR at various stages of recovery [17]. Within thirty seconds of exercise cessation, PSNS output rises rapidly and is reflective of cardiovascular autonomic function [17], peaking at approximately 2 minutes. The HR and BP therefore drop significantly within the first 2 minutes and plateau. A suboptimal HR drop after 2 minutes increases cardiovascular mortality [18]. This is expected to be faster among younger people [19]. Reduced HRR has however also been reported among obese, otherwise healthy females [20]. Chronotropic index is the percentage of one’s heart rate reserve for age utilised during exercise [21,22]. Chronotropic incompetence occurs when the chronotropic index is less than 0.8, or 0.62 in patients taking beta blockers [23,24]. It gives an individualised age-based assessment of cardiovascular autonomic function. Chronotropic index alone better predicts autonomic dysfunction and cardiovascular risk than HRR, while derangements in both parameters confer significantly higher cardiovascular risk [23].

Psychosocial factors are more prevalent in younger populations [25]. Among health workers, factors contributing to psychosocial stress include shift systems (including night and weekend shifts), emergency and/or critical care duties, taking end-of-life care decisions and relaying such decisions to patients and relatives, and having regular inpatient and outpatient services [26,27]. Few studies have reported the psychosocial factors experienced by junior doctors in Ghana, and even less is known about their association with changes in cardiovascular physiology. There is therefore a potential risk to this population that is uncharacterised. Understanding the psychosocial profile of junior doctors is relevant for developing interventions that address their overall wellbeing which will, in turn, improve their quality of life and healthcare provision.

This study therefore sought to determine the burden of depressive, anxiety and stress symptoms among junior doctors at the Korle-Bu Teaching Hospital (KBTH) and explore how these relate with their cardiovascular autonomic function, measured primarily by post-exercise heart rate recovery after one (1) minute (HRR_1_) and chronotropic index, and secondarily by heart rate recovery after two (2) minutes (HRR_2_)

## Materials and methods

The study was conducted at the Korle-Bu Teaching Hospital, the premier teaching hospital and largest public referral facility in Ghana. It also serves as the largest facility for postgraduate medical training across all specialties in Ghana.

A cross-sectional study was conducted from 17^th^ of April to the 18^th^ of August 2023. Junior doctors from departments with predisposing factors to psychosocial stress described earlier were considered for this study. Therefore, the selected departments were Internal Medicine, Surgery, Child Health, Obstetrics and Gynaecology, Anaesthesia, Emergency Medicine, and Family Medicine. These doctors had been at post for at least 6 months prior to the study. Exclusion criteria, due to potential effects on cardiovascular autonomic function, were:

- Pregnancy
- Self-declared previous or current diagnosis of hypertension, diabetes mellitus, hypothyroidism, hyperthyroidism, heart failure, chronic kidney disease or psychiatric illness.
- Doctors on beta blockers, renin-angiotensin-aldosterone system (RAAS) blockers or psychotropic medications
- Doctors with absolute contraindications for the TMT, e.g., inability to exercise, severe aortic stenosis, left bundle branch block on baseline electrocardiogram, etc.

### Sample size determination and sampling

The minimum sample size calculated using the Cochran formula at 95% confidence [28] and adjusting for a finite population of 864 junior doctors in the departments selected for this study was 241. With an anticipated 10% non-response rate, this was adjusted to 268. Though there is a paucity of data on stress, anxiety and depression among doctors in the West African sub-region, an Ethiopian study determined a 68.2% prevalence of stress among healthcare workers [26]. This gave a larger sample size than that obtained when calculated using a pooled prevalence of depression (29%) among doctors [29]. Participants were sampled using proportionate stratified sampling techniques based on department, rank and gender.

Lists of junior doctors were obtained from each selected department’s secretariats per rank in random order. Names of doctors in each rank were extracted, categorised based on gender and assigned serial numbers. The total number of doctors to be recruited per rank in each department was then determined proportionately based on gender. Subsequently, a sampling frame was applied based on the numbers of junior doctors in the departments per rank and sex, with the starting point determined by ballot.

### Operational definitions

House officer – A doctor who has completed medical school and is enrolled in the mandatory 2-year internship in 4 selected core rotations prior to permanent registration with the Medical and Dental Council of Ghana.

Junior doctor – Medical doctor who has not completed fellowship training in a medical/surgical specialty under a recognised postgraduate medical college.

Junior resident – A doctor enrolled in a postgraduate clinical training programme leading to the award of Membership of a postgraduate medical college.

Medical officer – A doctor who has completed the mandatory 2-year housemanship programme, is permanently licensed with the Ghana Medical and Dental Council but not enrolled in a residency training programme with a recognised postgraduate medical college.

Senior resident: A specialist who has enrolled in a Fellowship programme with a recognised postgraduate medical college.

Specialist – A doctor who has completed a clinical Membership programme with a recognised postgraduate medical college. They share the same professional responsibilities as a senior resident.

### Study Instruments

Data was obtained using a semi-structured questionnaire, anthropometric tools and the TMT. The questionnaire collected data on demographics, professional history, and psychosocial symptoms. The DASS-21 questionnaire was incorporated in the semi-structured questionnaire for the study. This was validated by Coker, Coker and Sanni in a Nigerian study [30]. The questionnaire was pre-tested on 10 junior doctors sampled from the various ranks using the same procedure intended for the study.

Weight was measured with a standard scale, measuring in kilograms (kg) to the nearest 0.1kg. A locally manufactured wooden stadiometer was used to measure height and subsequently converted from centimetres (cm) to metres (m), measured to the nearest 0.01m.

The TMTs were performed in line with recommendations on the conduct of TMTs by Smarz *et al.* [31], with a setup for resuscitation available if required. These were done during working hours with flexibility to accommodate varied work schedules (between 9am and 2pm) to ensure physiologic conditions were similar to what participants experience on a regular day.

### Data collection procedure

Selected participants were called on phone a week before the stress ECG to introduce the study. Consequently, the electronic questionnaire with an attached consent form detailing the study and unique serial number were forwarded via WhatsApp or electronic mail (e-mail) based on the participant’s preference. Regular reminders to complete the questionnaire were sent till the end of the following week. The TMT was scheduled once the questionnaire had been completed. The first question on the electronic questionnaire established consent to participate, while the second detailed the exclusion criteria. These were compulsory questions which required a “yes” and “no” response respectively to be allowed to participate. Any answers other than these rendered the participant disqualified. All those who met exclusion criteria were made to discontinue the study, and a replacement of the same rank, sex and department was selected randomly by ballot. Those who declined to fill the questionnaire were termed non-responders, as were those who had not filled the questionnaire by the end of the overall data collection period despite repeated reminders.

On arrival for the TMT, exclusion criteria were re-assessed to ensure qualification to participate. Anthropometric data were then obtained. Participants were ushered to a couch where they rested for 5 minutes, and vital signs measured. A portable electrocardiogram (Norav PC-ECG-1200 device) was fixed using the modified Mason-Likar technique and wirelessly connected to a console for continuous ECG monitoring. The baseline ECG was studied for ECG-based exclusion criteria. A symptom-limited Bruce protocol TMT was then performed, with no post-exercise cool-down stage. Indications for terminating the test included fatigue, limiting chest pain, light-headedness, leg cramping, ventricular tachycardia, new-onset ST segment elevations more than 1mm above baseline ECG, systolic BP drop by 10mmHg during exercise, and participant’s request to discontinue for other reasons. Participants who were unfamiliar with the use of the treadmill had a 1-minute warm-up session at speeds from 0.5km/h up to 1.5km/h, to allow them to adjust to its use prior to commencing the Bruce protocol. On completion of the exercise stages, participants had a 3-minute recovery stage.

### Data management

Autonomic function assessed by HRR_1_ and chronotropic index from the TMT was the primary outcome. As a secondary outcome, HRR_2_ was also assessed.

The maximal predicted heart rate (MPHR) was defined as “220-age in years” [31].

HRR_1_ was determined by “heart rate at peak exercise minus heart rate after the 1^st^ minute of recovery”. HRR_1_ values ≤ 18bpm were considered abnormal [18,32,33]. All heart rate determinations were based on ECG tracings from the wireless electrocardiographic device.

The chronotropic index was calculated as (peak HR-resting HR) / (maximum predicted HR – resting HR). Chronotropic incompetence was defined as an index < 0.80 [23].

HRR_2_ was calculated as the difference between HR at peak exercise minus HR at 2 minutes of recovery, with values of ≤42bpm considered abnormal [22].

### Psychosocial assessment

Scores obtained from answers to questions for depressive, anxiety and stress symptom categories from the DASS-21 scale were summed and doubled. Consequently, they were used to determine the classification of the respective symptoms as shown in Table 1 below.

**Table 1.** Categorisation of depressive, anxiety and stress symptoms according to the DASS-21 scale.

|  | Depression | Anxiety | Stress |
| --- | --- | --- | --- |
| Normal | 0-9 | 0-7 | 0-14 |
| Mild | 10-13 | 8-9 | 15-18 |
| Moderate | 14-20 | 10-14 | 19-25 |
| Severe | 21-27 | 15-19 | 26-33 |
| Extremely severe | 28+ | 20+ | 34+ |

### Statistical analysis

The completed questionnaire was exported from Google Forms to a Microsoft Excel spreadsheet. The dataset was then exported, cleaned, coded, and analysed using Stata statistical software version 16.1. Missing responses within the DASS-21 subscale were handled using person mean substitution, since at most one item within the respective subscale was missing. Continuous variables were summarized using mean and standard deviation (SD) for approximately normally distributed data, and median with interquartile range (IQR) for skewed distributions. Categorical variables were summarized as frequencies and percentages. Internal consistency of the DASS-21 subscales was assessed using Cronbach’s alpha, while sampling adequacy was evaluated with the Kaiser-Meyer-Olkin (KMO) measure.

At the bivariate level, correlations among DASS-21 subscales, between each DASS-21 subscale and cardiovascular autonomic function measures, namely chronotropic index and HRR1, and between the two autonomic outcomes were assessed. Pearson correlation coefficients were used for normally distributed variable pairs, while Spearman’s rank correlation was applied for nonparametric data. Differences in chronotropic index and HRR1 across tertiles of each DASS-21 subscale were evaluated using one-way analysis of variance (ANOVA) for normally distributed outcomes or the Kruskal-Wallis test for skewed outcomes. Corresponding box plots were generated for graphical presentation.

For multivariable analysis, DASS-21 subscale scores were standardized to a mean of 0 and a standard deviation of 1 to facilitate interpretable effect sizes relating to the chronotropic index and HRR1. Primary analyses were conducted using linear regression models, adjusting for biologically relevant covariates, following assessment of residuals and other model assumptions. Quantile regression was fitted for the chronotropic index model due to violations of multiple linear regression diagnostics. For HRR1, linear regression with robust variance estimation was used to account for the presence of outliers. Secondary analyses employed multivariable logistic regression models for categorized chronotropic index and HRR1 based on predefined clinical cutoffs. Linearity in the logit for continuous predictors was assessed graphically and found to be satisfactory; thus, predictors were retained in their original continuous form. Associations were expressed per standard deviation increase in DASS-21 scores to facilitate clinically interpretable effect sizes and comparison across psychosocial constructs. Statistical significance was set at a two-sided p-value less than 0.05, with a corrected threshold of 0.01 to account for multiple comparisons involving two co-primary outcomes and three factor variables.

### Ethical considerations

Approval was obtained from the Institutional Review Board (IRB) of the Korle-Bu Teaching Hospital (REF No. KBTH-IRB 000154/22). Written informed consent was obtained from each participant. Study data were completely anonymised.

## Results

### Demographic characteristics of respondents

A total of 268 junior doctors were invited to participate in this study, with a response rate of 91% (244/268). Sociodemographic and clinical characteristics are shown in Table 2. Among respondents, 123 (50.41%) were male. The mean age of participants was 31.46 (SD 4.41) years. The median body mass index (BMI) was 25.58 (IQR, 22.79-28.66) kg/m^2^. Sixty-nine (69) participants reported that they consume alcoholic beverages (28.28%) at least occasionally, while 6 (2.46%) and 100 (40.98%) reported smoking (tobacco and shisha) and drinking coffee, respectively.

**Table 2.** Sociodemographic and clinical characteristics of study participants (N = 244)

| Characteristic | Value (%) |
| --- | --- |
| *Sex* |  |
| Male | 123 (50.41%) |
| Female | 121 (49.59%) |
| *Age (years), mean (SD)* | 31.46 (4.41) |
| ***Age-group*** |  |
| 25 years and below | 26 (10.66%) |
| 26-30 years | 81 (33.20%) |
| 31-35 years | 86 (35.25%) |
| 36-40 years | 45 (18.44%) |
| 41-45 years | 5 (2.05%) |
| Above 45 years | 1 (0.41%) |
| ***Marital status*** |  |
| Single | 128 (52.46%) |
| Married | 114 (46.72%) |
| Divorced | 2 (0.82%) |
| ***Years worked as a doctor*** |  |
| Less than 2 years | 66 (27.05%) |
| 2-5 years | 67 (27.46%) |
| 5-10 years | 89 (36.48%) |
| More than 10 years | 22 (9.02%) |
| ***Grade/Rank*** |  |
| House officer | 65 (26.64%) |
| Medical officer | 59 (24.18%) |
| Junior resident | 85 (34.84%) |
| Senior resident | 35 (14.34%) |
| ***Years in current rank*** |  |
| Less than 2 years | 162 (66.39%) |
| 2-5 years | 74 (30.33%) |
| 5-10 years | 7 (2.87%) |
| More than 10 years | 1 (0.41%) |
| ***Department*** |  |
| Anaesthesia | 33 (13.52%) |
| Child health | 36 (14.75%) |
| Emergency medicine | 18 (7.38%) |
| Family medicine | 19 (7.79%) |
| Internal medicine | 60 (24.59%) |
| Obstetrics and Gynaecology | 33 (13.52%) |
| Surgery | 45 (18.44%) |
| ***Average hours of work per week*** |  |
| Less than 30 hours | 4 (1.64%) |
| 30 – 39 hours | 65 (26.64%) |
| 40 – 49 hours | 85 (34.84%) |
| 50 hours or more | 90 (36.89%) |
| ***Alcohol consumption*** |  |
| Yes | 69 (28.28%) |
| No | 175 (71.72%) |
| ***Smoking*** |  |
| Yes | 6 (2.46%) |
| No | 238 (97.54%) |
| ***Coffee intake*** |  |
| Yes | 100 (40.98%) |
| No | 144 (59.02%) |
| ***BMI (kg/m<sup>2</sup>), median (IQR)*** | 25.58 (22.79-28.66) |

Values are presented as frequency (percentage), mean (SD), or median (IQR).
Abbreviations: BMI, body mass index; IQR, interquartile range; kg/m<sup>2</sup>, kilogram per meter square; N, sample size; SD, standard deviation

### Psychosocial factor assessment

The DASS-21 demonstrated good internal consistency, with Cronbach’s alpha values of 0.86, 0.80, and 0.90 for the stress, anxiety, and depression subscales, respectively. The Kaiser-Meyer-Olkin (KMO) measure indicated excellent sampling adequacy (overall KMO = 0.92). The distribution of DASS-21 subscale scores is shown in Table 3. Stress scores were approximately normally distributed, with a mean score of 15.51 (SD 8.65), whereas anxiety and depression scores were skewed, with median anxiety and depression scores of 6 (IQR 2-14) and 4 (IQR 0-10), respectively. The prevalence of stress, anxiety, and depressive symptoms was determined to be 48.36%, 48.77%, and 29.51%, respectively, with varied degrees of symptom severity (i.e., from mild to extremely severe). Participants were relatively evenly distributed across low, middle, and high score tertiles for the stress subscale, while they were largely in low tertiles for anxiety and depression subscales.

**Table 3.** Distribution of DASS-21 subscale scores among study participants (N=244)

| Variable | Stress | Anxiety | Depression |
| --- | --- | --- | --- |
| Mean/median score | 15.51 (SD 8.65) | 6 (IQR 2-14) | 4 (IQR 0-10) |
| Severity category, n (%) |  |  |  |
| Normal | 126 (51.64) | 125 (51.23) | 172 (70.49) |
| Mild | 35 (14.34) | 14 (5.74) | 23 (9.43) |
| Moderate | 50 (20.49) | 50 (20.49) | 31 (12.70) |
| Severe | 26 (10.66) | 28 (11.48) | 13 (5.33) |
| Extremely severe | 7 (2.87) | 27 (11.07) | 5 (2.05) |
| Tertiles□, n (%) |  |  |  |
| Low | 93 (38.11) | 102 (41.80) | 106 (43.44) |
| Middle | 84 (34.43) | 74 (30.33) | 66 (27.05) |
| High | 67 (27.46) | 68 (27.87) | 72 (29.51) |

Scores are presented as mean (SD) for normally distributed variables (stress) and median (IQR) for skewed variables (anxiety and depression). Categorical variables are presented as a number, n (percentage, %). Severity categories are based on DASS-21 subscale cut-offs. Abbreviations and symbols: n, number; SD, standard deviation; IQR, interquartile range. □ Tertiles of the respective subscale scores.

There was a moderate positive correlation between stress and anxiety scores (Spearman’s ρ = 0.55, p < 0.001), stress and depression scores (Spearman’s ρ = 0.61, p < 0.001), and anxiety and depression scores (Spearman’s ρ = 0.57, p < 0.001).

### Treadmill stress test and cardiovascular autonomic function parameters

All participants underwent a symptom-limited TMT using the standard Bruce protocol. Fatigue was the main reason for TMT termination (68.9%). The median resting heart rate was 81 (IQR, 73-88) bpm, while the median peak heart rate was 175 (IQR, 165 – 180.5) bpm. Up to 238 participants (97.54%) achieved the target heart rate, i.e., at least 85% of the maximum predicted heart rate (median, 94.5%; IQR, 90%-100%).

For the cardiovascular autonomic function measures (Table 4), the HRR_1_ and HRR_2_ were approximately normally distributed, and the mean HRR_1_ was 38.09 (SD, 9.45; 95% CI, 36.9-39.29) bpm, while the mean HRR_2_ was 58.14 (SD, 9.82; 95% CI, 56.90-59.38) bpm. Chronotropic index data were skewed, with a median of 0.92 (IQR, 0.85 – 1.00; bootstrap 95% CI, 0.89-0.94). While an abnormal chronotropic index (chronotropic incompetence) was observed in 23 participants (9.43%; 95% CI, 6.07% - 13.81%), HRR_1_ and HRR_2_ were abnormal among 8 (3.28%; 95% CI, 1.43% - 6.36%) and 15 (6.15%; 95% CI, 3.48% −9.94%) participants, respectively.

**Table 4.**
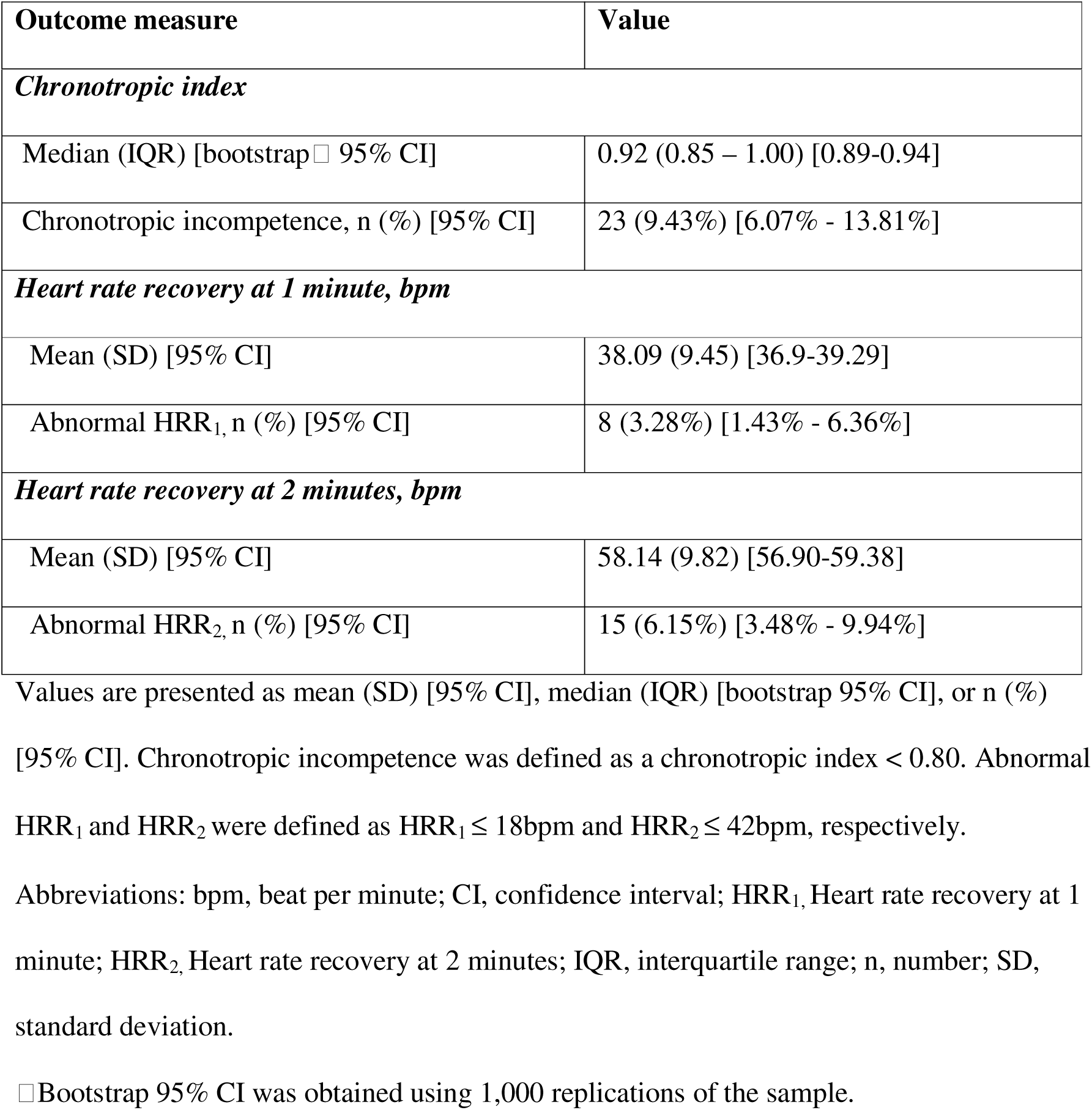
Summary and categorical distribution of cardiovascular autonomic function measures among study participants (N = 244)

| Outcome measure | Value |
| --- | --- |
| ***Chronotropic index*** |  |
| Median (IQR) [bootstrap □ 95% CI] | 0.92 (0.85 – 1.00) [0.89-0.94] |
| Chronotropic incompetence, n (%) [95% CI] | 23 (9.43%) [6.07% - 13.81%] |
| ***Heart rate recovery at 1 minute, bpm*** |  |
| Mean (SD) [95% CI] | 38.09 (9.45) [36.9-39.29] |
| Abnormal HRR<sub>1</sub>, n (%) [95% CI] | 8 (3.28%) [1.43% - 6.36%] |
| ***Heart rate recovery at 2 minutes, bpm*** |  |
| Mean (SD) [95% CI] | 58.14 (9.82) [56.90-59.38] |
| Abnormal HRR<sub>2</sub>, n (%) [95% CI] | 15 (6.15%) [3.48% - 9.94%] |

Values are presented as mean (SD) [95% CI], median (IQR) [bootstrap 95% CI], or n (%) [95% CI]. Chronotropic incompetence was defined as a chronotropic index < 0.80. Abnormal HRR<sub>1</sub> and HRR<sub>2</sub> were defined as HRR<sub>1</sub> ≤ 18bpm and HRR<sub>2</sub> ≤ 42bpm, respectively. Abbreviations: bpm, beat per minute; CI, confidence interval; HRR<sub>1</sub>, Heart rate recovery at 1 minute; HRR<sub>2</sub>, Heart rate recovery at 2 minutes; IQR, interquartile range; n, number; SD, standard deviation.
□ Bootstrap 95% CI was obtained using 1,000 replications of the sample.

Chronotropic index and HRR_1_ were not correlated (ρ = −0.09, p = 0.17), while HRR_1_ and HRR_2_ were strongly correlated (Pearson’s r = 0.77, p < 0.001).

### Relationship between psychosocial factors and cardiovascular autonomic function

Chronotropic index was not correlated with DASS-21 stress (ρ = −0.09, p = 0.15); anxiety (ρ = −0.05, p = 0.45); or depression (ρ = 0.00, p = 0.96). Similarly, HRR_1_ was not correlated with DASS-21 stress (r = 0.11, p = 0.09); anxiety (ρ = 0.03, p = 0.64); or depression (ρ = - 0.01, p = 0.90).

Box plots showed similar distributions of both chronotropic index and HRR_1_ across tertiles of stress, anxiety, and depression scores, with largely overlapping medians and interquartile ranges (Figs 3a-c and 4a-c). The Kruskal-Wallis test revealed no difference in chronotropic index across tertiles of stress (χ² (2) = 3.89, p = 0.14); anxiety (χ² (2) = 1.12, p = 0.57); or depression (χ² (2) = 0.03, p = 0.99) (S1 Table). Similarly, one-way ANOVA showed no difference in mean HRR_1_ across tertiles of stress (F_2,241_ = 1.90, p = 0.15); anxiety (F_2,241_ = 0.37, p = 0.69); or depression (F_2,241_ = 0.05, p = 0.95) (S2 Table).

**Fig 3a-c.**
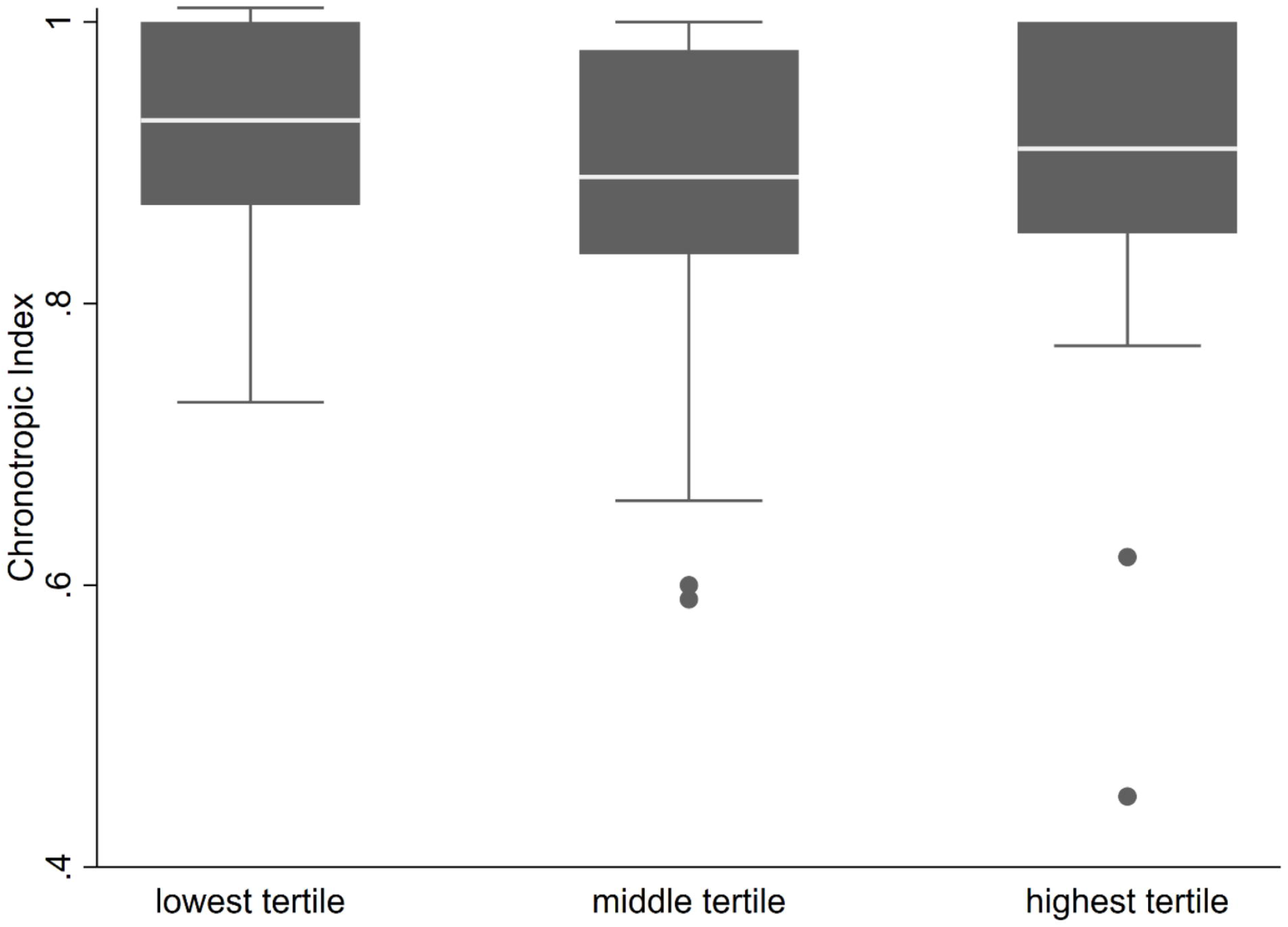
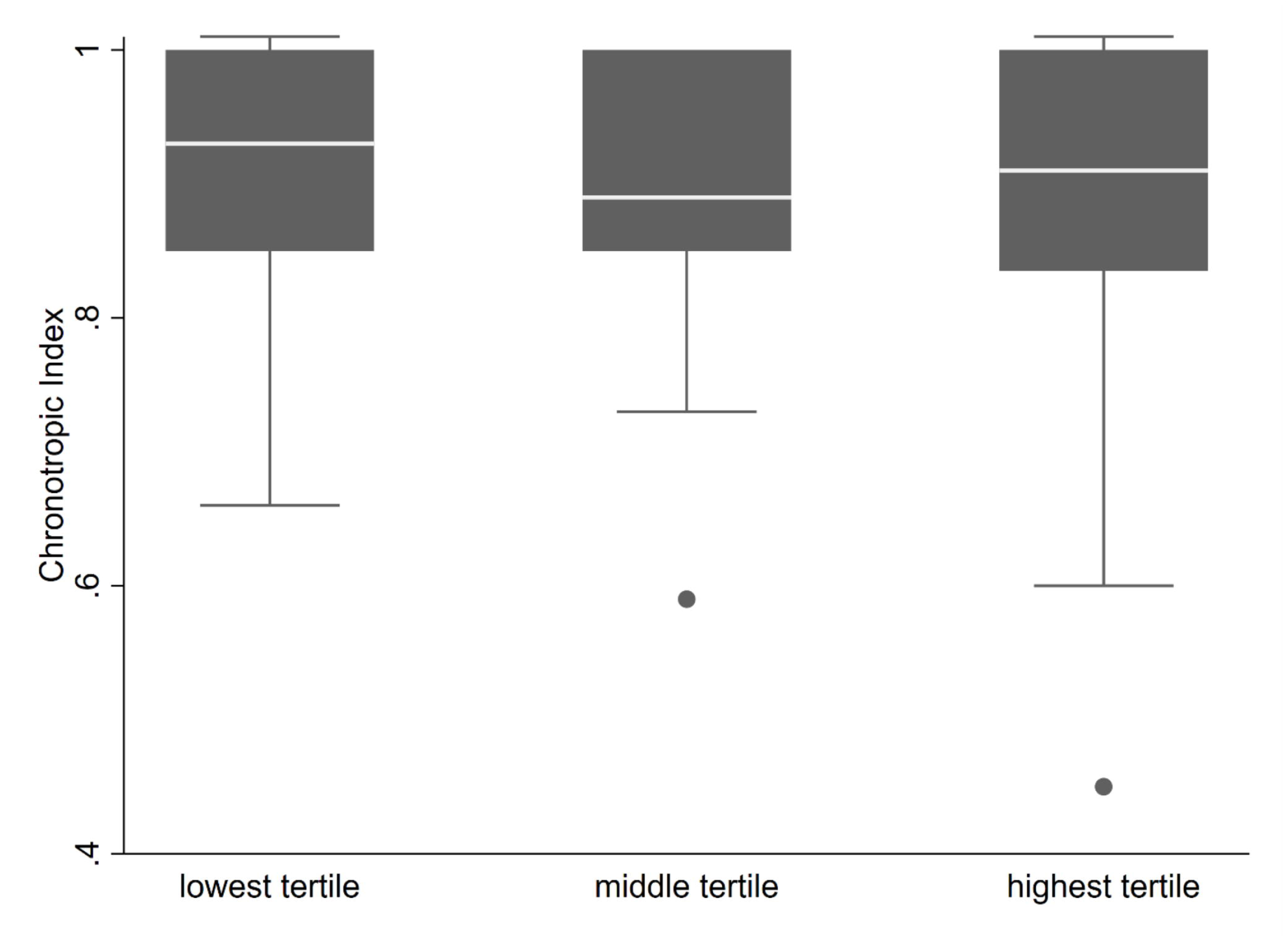
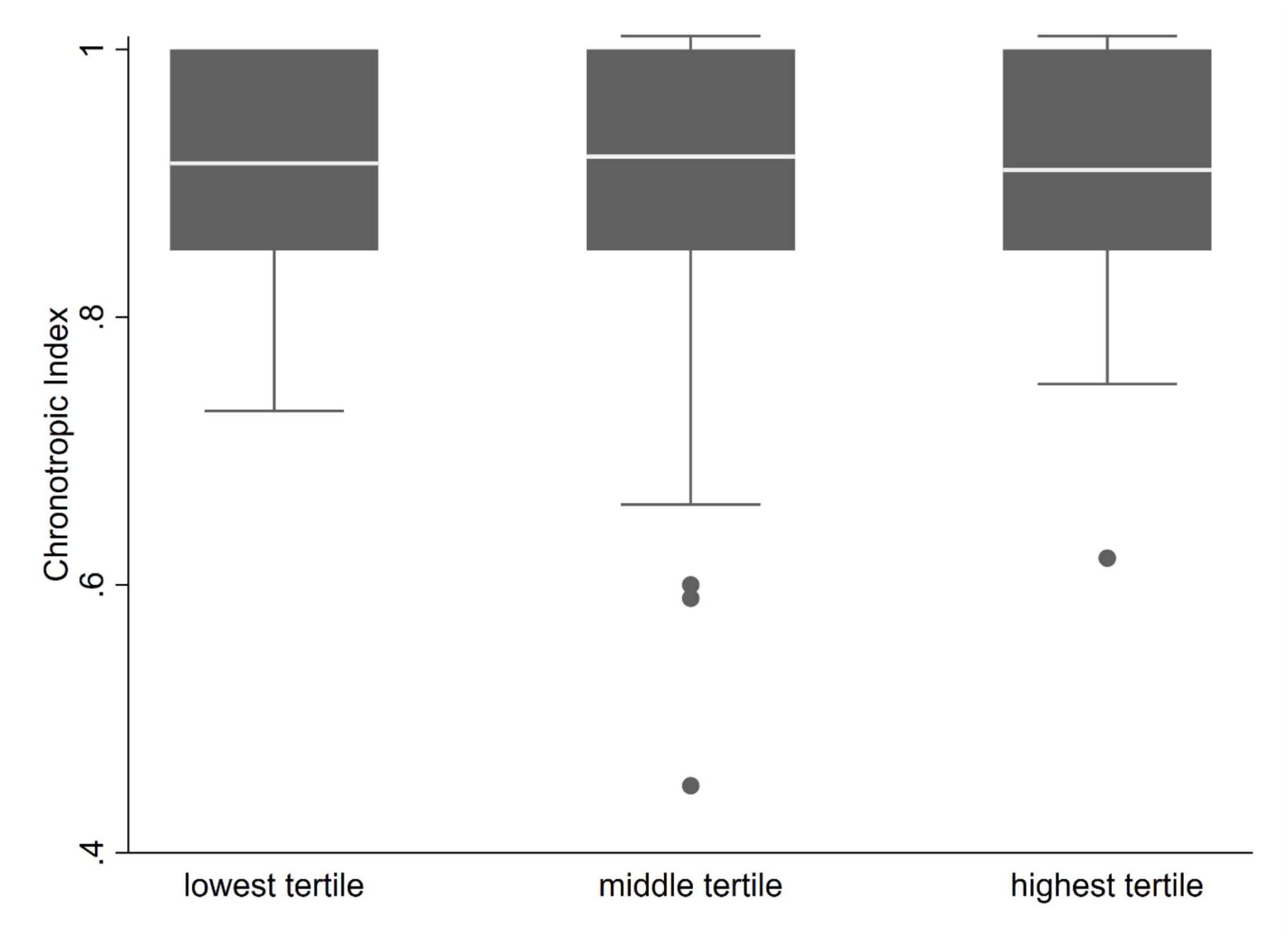
Box and whisker plots of chronotropic index across tertiles of psychosocial factor subscales assessing group differences using the Kruskal-Wallis test. Boxes represent interquartile ranges with median lines, and whiskers indicate 1.5 times the interquartile range. 3a, chronotropic index vs stress tertiles, χ² (2) = 3.89, p = 0.14. 3b) chronotropic index vs anxiety tertiles, χ² (2) = 1.12, p = 0.57. 3c) chronotropic index vs depression tertiles, χ² (2) = 0.03, p = 0.99.

**Figure 4a-c.**
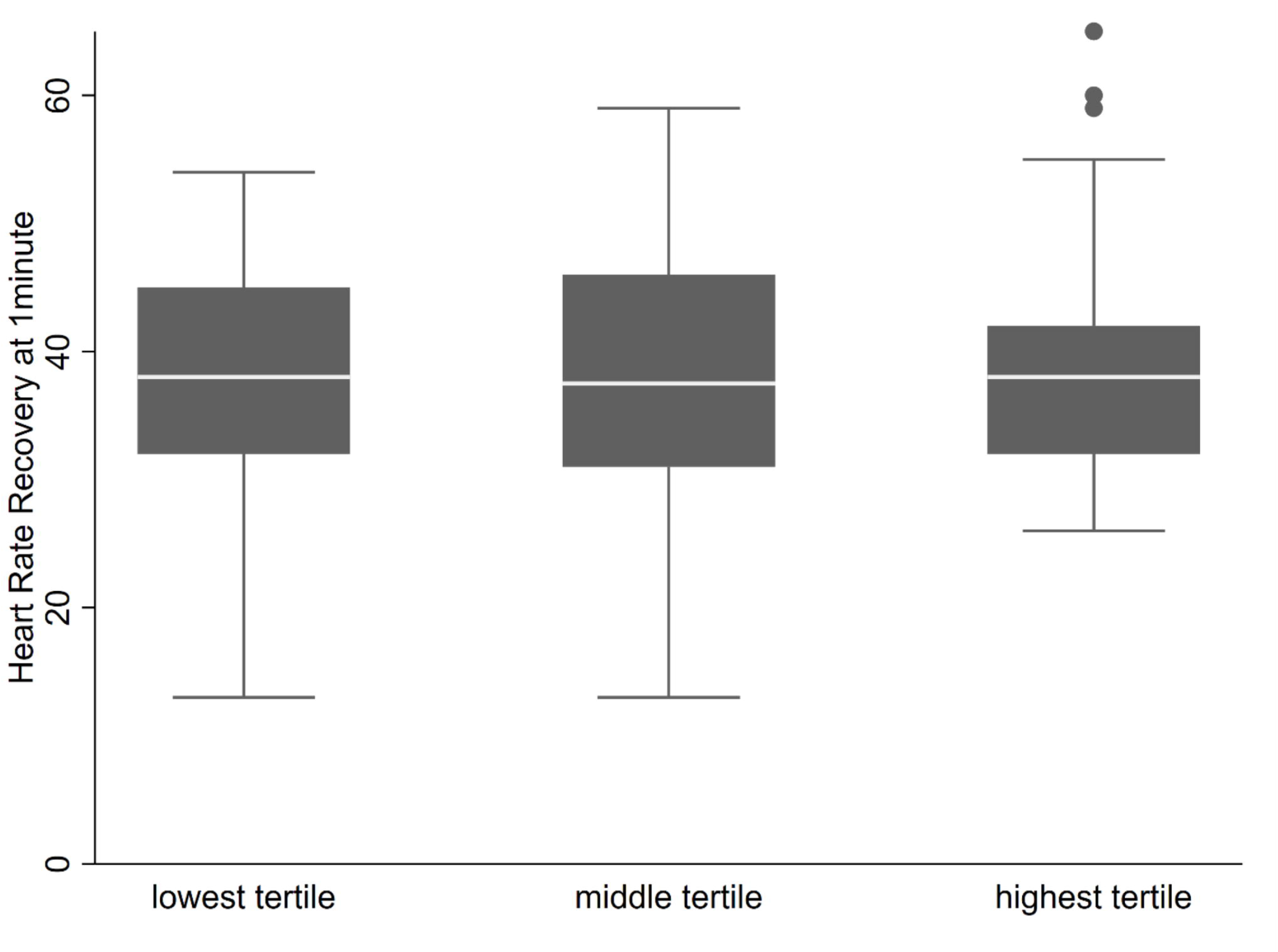
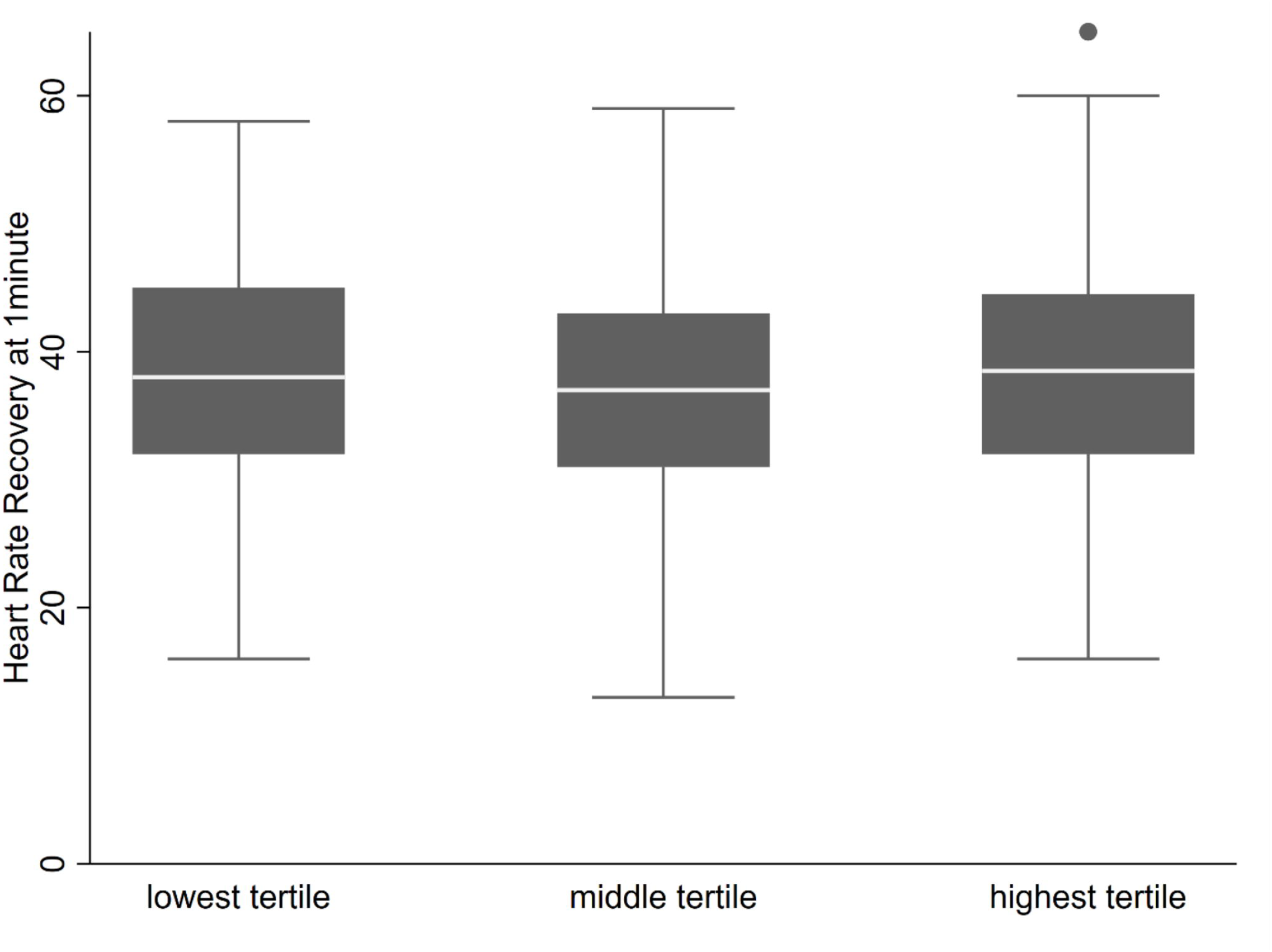
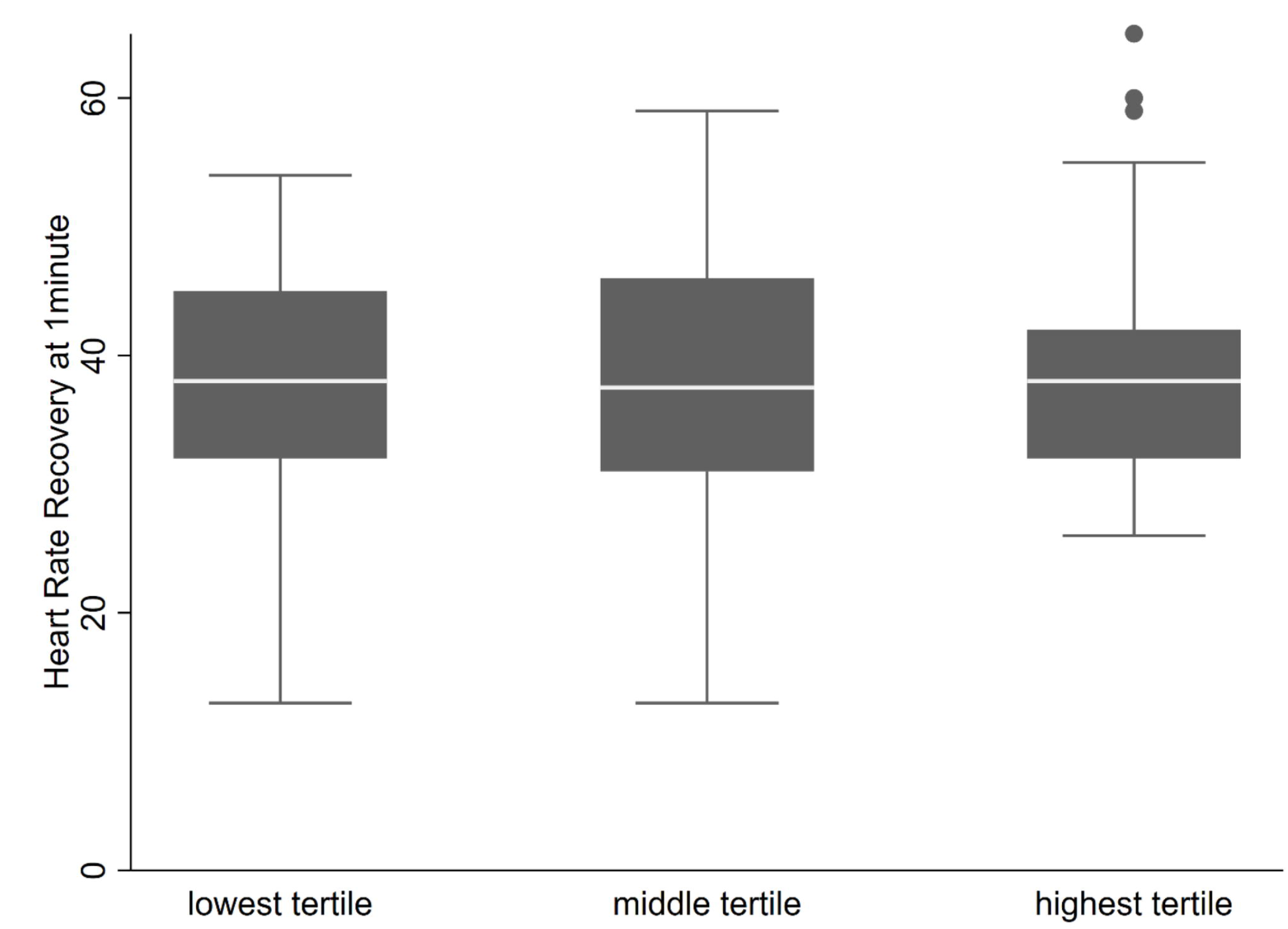
Box and whisker plot of heart rate recovery at 1 minute (HRR_1_) across tertiles of psychosocial factor subscales assessing group differences using one-way ANOVA. Boxes represent interquartile ranges with median lines, and whiskers indicate 1.5 times the interquartile range. 4a), HRR_1_ vs stress tertiles, F (2, 241) = 1.90, p = 0.15. 4b) HRR_1_ vs anxiety tertiles, F (2, 241) = 0.37, p = 0.69. 4c) HRR_1_ vs depression tertiles, F (2, 241) = 0.05, p = 0.95.

After adjustment for age, sex, BMI, smoking, coffee intake, and resting heart rate, higher DASS-21 stress scores were associated with a small reduction in chronotropic index at the median in quantile regression, although the 95% CI included the null (β per SD −0.01, 95% CI −0.03 - 0.00; Table 5). Anxiety and depression scores showed minimal associations with chronotropic index, with effect estimates close to zero (S3-6 Tables). In contrast, higher stress scores were associated with higher HRR1 in adjusted linear regression (β per SD 1.53 bpm, 95% CI 0.03 - 3.02), while anxiety and depression scores were not meaningfully associated with HRR1 (Table 5). In logistic regression using clinically defined cut-offs (S7-10 Tables), higher stress and anxiety scores were associated with increased odds of chronotropic incompetence, although estimates were imprecise (adjusted OR per stress score SD 1.68, 95% CI 0.93 - 3.04; adjusted OR per anxiety score SD 1.30, 95% CI 0.73 – 2.31), whereas higher depression scores were associated with lower odds of chronotropic incompetence (adjusted OR 0.44, 95% CI 0.22 - 0.88; Table 6). Associations between DASS-21 subscales and abnormal HRR1 were generally weak and characterized by wide confidence intervals spanning clinically relevant effects in both directions (Table 6).

**Table 5.** Quantile and linear regression models assessing the association between psychosocial factors and autonomic function.

| DASS-21<br>subscale | Unadjusted coefficient, $\beta$(95% CI)<br>[p-value] | Adjusted coefficient, $\beta$(95%<br>CI) [p-value] |
| --- | --- | --- |
| *Chronotropic index: Quantile regression, n= 244, Pseudo R<sup>2</sup>= 0.12* |  |  |
| Stress score | $-0.02$ ($-0.03 - -0.00$) [$0.03$] | $-0.01$ ($-0.03 - 0.00$) [$0.17$] |
| Anxiety score | $-0.01$ ($-0.03 - 0.01$) [$0.19$] | $-0.00$ ($-0.02 - 0.01$) [$0.68$] |
| Depression score | -0.00 (-0.02 – 0.01) [0.62] | 0.01 (-0.01 – 0.02) [0.50] |
| *HRR<sub>1</sub>: Linear regression with vce robust, n=244, R<sup>2</sup>=0.11, p=0.03* |  |  |
| Stress score | 1.01 (-0.18 – 2.20)bpm [0.09] | 1.53 (0.03 – 3.02)bpm [0.04] |
| Anxiety score | 0.40 (-0.76 – 1.57)bpm [0.49] | -0.08 (-1.46 – 1.31)bpm [0.91] |
| Depression score | 0.14 (-0.90 – 1.18)bpm [0.78] | -0.60 (-1.82 – 0.61)bpm [0.33] |

Chronotropic index was modelled using quantile regression at the median, while HRR<sub>1</sub> was analysed using linear regression with robust variance estimation (vce). Regression coefficients ( $\beta$ ) represent effects per one standard deviation increase in the respective DASS-21 subscale score. Adjusted models included age, sex, body mass index, smoking status, coffee intake, and resting heart rate. Results are presented with 95% confidence intervals to emphasize effect size and precision; p-values are provided for completeness. Abbreviations: bpm, beats per minute; CI, confidence interval; HRR<sub>1</sub>, Heart rate recovery at 1 minute; n, sample; R<sup>2</sup>, R-squared of the model.

**Table 6.**
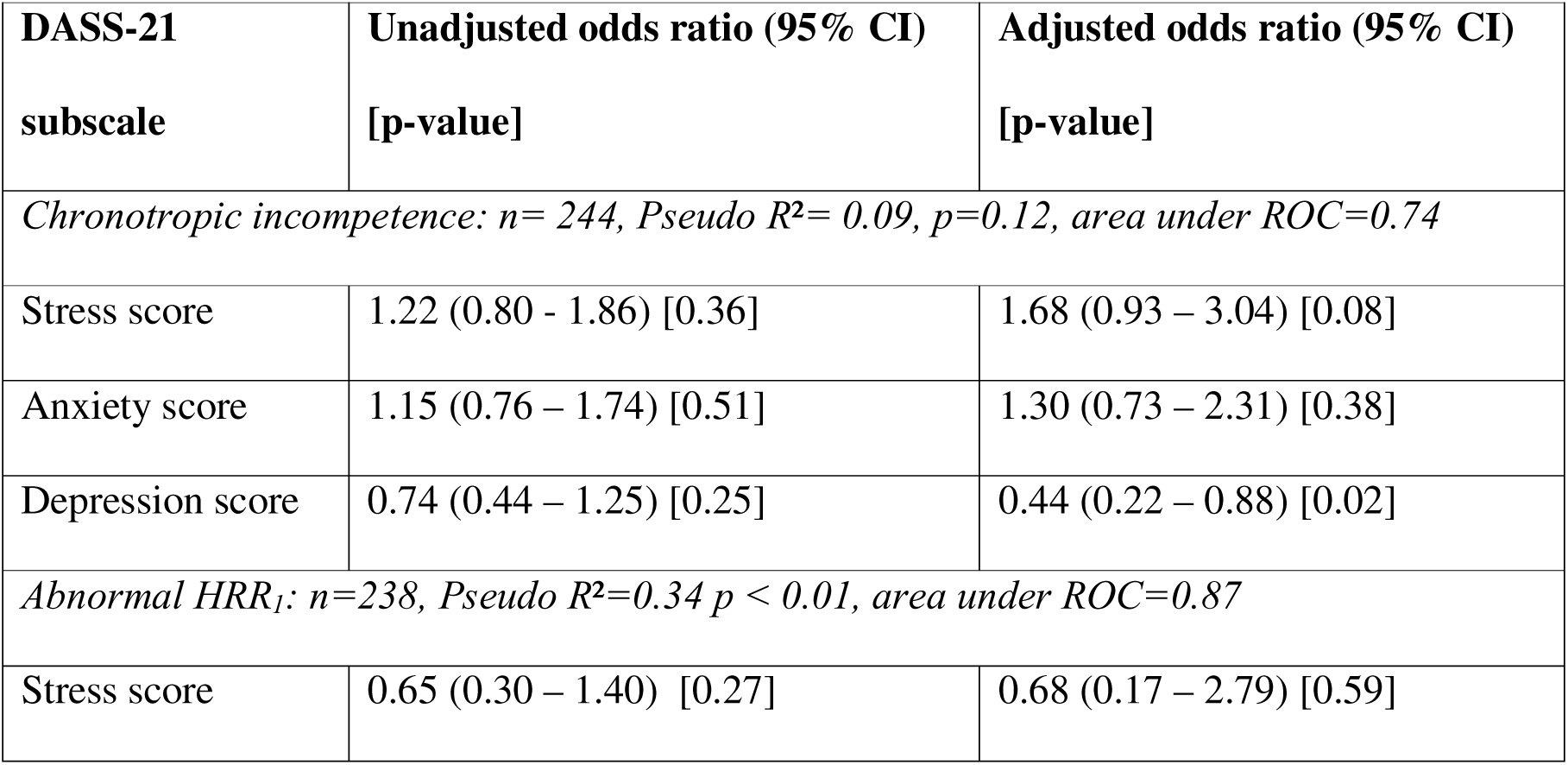
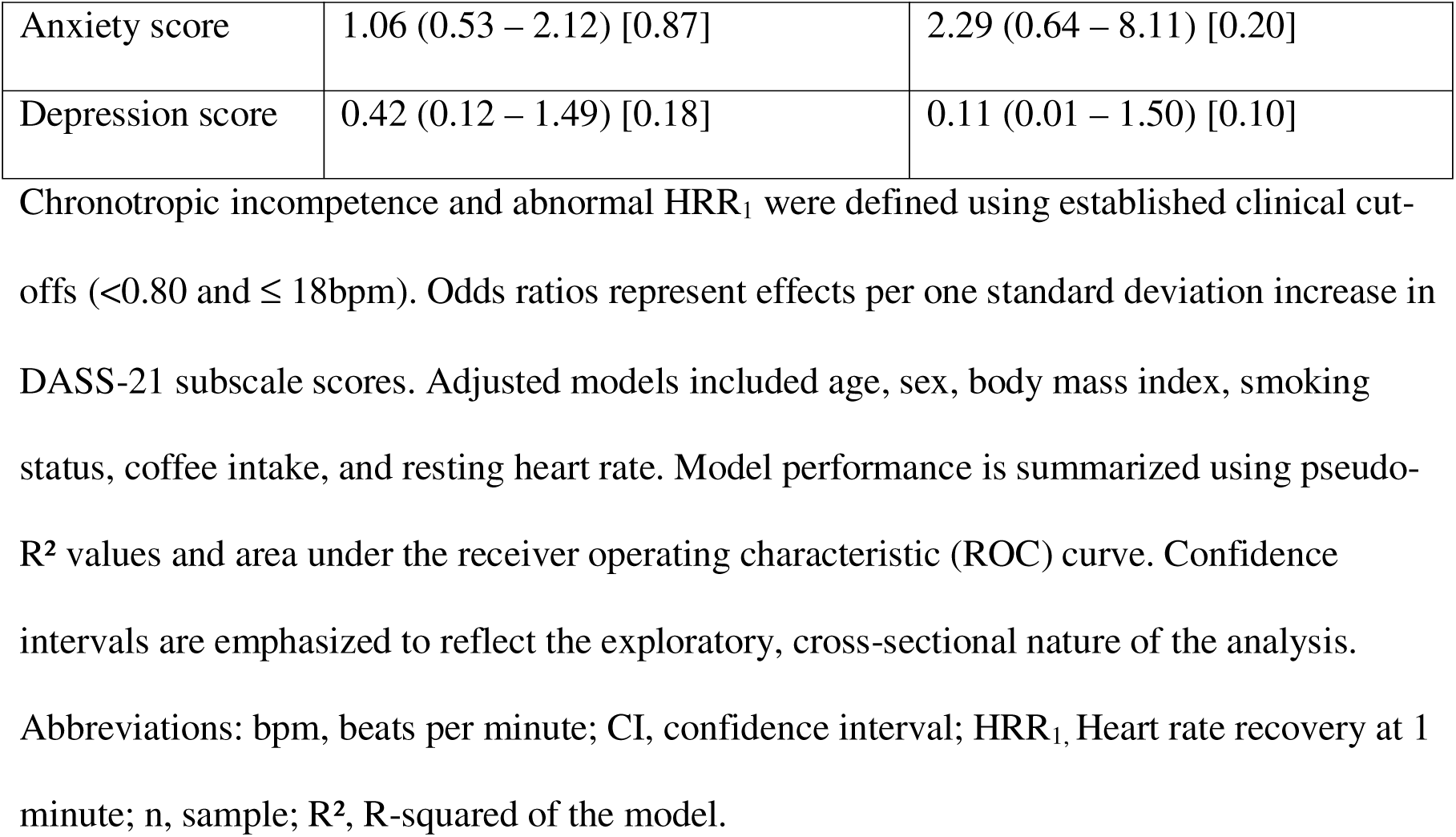
Logistic regression models assessing the association between psychosocial factors and autonomic function.

## Discussion

The study was conducted among junior doctors at the KBTH, with a mean age of 31.46 years. It revealed a high prevalence of psychosocial factors which correlated moderately with one another. Less than 10% of participants recorded abnormal parameters of cardiovascular autonomic function, with no significant differences after comparing based on tertiles of the psychosocial subscales. There was no relationship between psychosocial factors and chronotropic index after adjusting for possible confounders, while higher stress scores were associated with an increase in HRR_1_ in a linear regression model. High depression scores were however associated with decreased odds of chronotropic incompetence.

The high KMO and Cronbach’s alpha across all psychosocial subscales assure adequate sampling and internal consistency of the DASS-21 tool within the sample population. The prevalences of stress, anxiety and depression were 48.36%, 48.77% and 29.15% respectively.

These are consistent with other studies, with a systematic review by Mata et al. reporting a 28.8% pooled prevalence of depression [29]. Early stages of professional development have been associated with higher rates of psychosocial distress [34], consistent with the population demographics of this study. Despite the use of different stress assessment scales, studies in Ethiopia [26,35] and Saudi Arabia [36] all showed high levels of stress, ranging from 48-73%. These factors also correlated with one another as already established [37], underscoring their intricate relationship and the concern for multimorbidity. Over 75% of this study’s population comprised doctors in training positions, suggesting multiple responsibilities; conducting daily inpatient reviews, taking responsibility for day-to-day patient care decisions, running multiple clinics under supervision, providing daily cover for wards and the emergency unit, seeking out opportunities to observe and practice procedural skills required to be proficient for the next career stage, and seeking to balance these with family and personal commitments. Similar factors were described as contributory to stress among Nigerian junior doctors [38].

Participants with moderate psychosocial symptoms or higher across all assessed factors represented majority of those who reported symptoms, consistent with findings from Saudi Arabia which show a 34.9% prevalence of severe stress among interns (house officers) [36]. Similarly, a 21% and 58% prevalence of at least moderate levels of stress were reported in studies among nurses and midwives in Northern Ghana respectively [39,40], in consonance with our findings. The presence of these symptoms may the risk of unforced work-related errors and the adoption of risky lifestyle behaviours [37,41] which may harm patients inadvertently, though it has been suggested that there is an inconsistent relationship between stress and risky behaviours such as alcohol use, smoking and physical inactivity [42]. Such levels of psychosocial stress could progress to burnout as suggested by studies among Ghanaian and Nigerian health workers [27,43]. Consequently, sufferers may experience downstream cardiovascular and psychological/psychiatric effects [4,34,37,44].

There was a low prevalence of cardiovascular autonomic dysfunction with no significant variation when analysed across tertiles of psychosocial factors. This could be due to high resilience because of good coping mechanisms, because while psychosocial factors invoke an autonomic response, the development of autonomic dysfunction follows long-term exposure to stressors that overwhelm intrinsic coping mechanisms [6,8]. Alternatively, there may be an asymptomatic stage that the tools used in this study are not sensitive to diagnose. Heart rate variability (HRV) has been used in assessing cardiovascular autonomic function in other studies, including a study among Ghanaian nurses which found no association with occupational stress [40]. However, a meta-analysis on the relationship between HRV and occupational stress identified 8 out of 10 studies across various professions that demonstrated abnormal cardiovascular autonomic function [45]. HRR is described by Goldberger et al. as greater in assessing the dynamic changes of the autonomic nervous system, though both HRR and HRV have limited real-world clinical utility [17]. Though a study with a small number of participants identified a significant correlation between HRR and HRV, other evidence comparing them have been equivocal, with neither showing a clear diagnostic advantage [46].

Chronotropic index showed no relationship with the independent variables when adjusted for confounding, whereas HRR_1_ values were higher with higher stress scores. The associated SNS activation in response to stress, together with the exercise-induced increase in SNS output, can result in higher peak HR following exercise. This could lead to marked HR reductions after exercise as the PSNS output increases, leading to increases in HRR. When PSNS output is however diminished due to chronic exposure to the psychosocial stressors, this may then manifest as autonomic dysfunction. As seen in Figure 1, it may be plausible that the study participants, despite the high prevalence of psychosocial factors, have not progressed to the chronic state responsible for autonomic dysfunction. Additionally, stressors may rather improve efficiency and resourcefulness based on one’s subjective assessment of the situation and coping mechanisms. This is called eustress [9,26]. This is consistent with the population mean and medians for the psychosocial constructs falling within the mild symptom bracket, suggesting that the stressors they face overall may not be overwhelming yet. Similar mechanisms could explain the lower odds of chronotropic incompetence related to depression scores.

The low prevalence of cardiovascular autonomic dysfunction may be attributable to the relatively young ages of the study population (mean age = 31.46 years), presumably shorter exposure to psychosocial stressors, coping mechanisms, or other factors not explored in this study. Additionally, doctors may be presumed to be more health conscious than the general population, putting effort into controlling psychosocial stressors. The individual response to stress is however varied and difficult to characterise distinctly [42]. The small effect sizes and wide confidence intervals of some outcome parameters may reflect limited statistical power to identify such associations. Nonetheless, the prevalences of depressive, anxiety and stress symptoms have clinical significance that requires attention to safeguard the health of the junior doctor. It is therefore recommended that the findings of this study be interpreted as exploratory, generating hypothesis for further longitudinal investigation. These can assess how psychosocial factors relate with cardiovascular autonomic function and potential mechanisms for this relationship, strengthened with the use of invasive psychosocial stress markers like cortisol and catecholamine levels to enhance objectivity.

## Limitations

While the study has some strengths, there are significant limitations that need to be acknowledged. The use of the DASS-21 tool is subject to recall/self-report bias, potentially resulting in an over- or under-estimation of the true burden of stress. Secondly, the duration of exposure to the psychosocial factors may not qualify as chronic, depending on the time over which this has happened and coping mechanisms. The definition of acute versus chronic stress is individualised based on each person’s threshold determined by one’s perception of situations and coping mechanisms. This could influence results as the longstanding haemodynamic changes from autonomic dysfunction are due to chronic stress, rather than exposure to stress acutely. Additionally, the study could not differentiate between eustress, which improves output, and distress which causes deleterious effects. Routine exercise habits, sleep patterns and shifts before exercise were not factored into the study, which could potentially have influenced the results as autonomic function was assessed. Finally, this was a single-centre cross-sectional study, limiting the generalisability of the results as well as the inference of causality.

## Conclusion

There are high prevalences of stress and anxiety symptoms among junior doctors at the KBTH, with a lower (albeit significant) prevalence of depressive symptoms. The prevalence of cardiovascular autonomic dysfunction was low, and some psychosocial factors were associated with enhanced cardiovascular autonomic function suggesting a more acute duration of these factors or durable coping mechanisms. There were no significant differences in cardiovascular autonomic function based on severity of psychosocial factors.

It is suggested that sources and effects of these psychosocial stressors be studied further with adequate sampling to enable improved efforts at mental health protection of the junior doctor.

## Acknowledgements

Dr Muhyideen Bashir assisted with the review of the statistical analysis and interpretation of results in the revised manuscript.

The efforts of Dorcas Baiden-Amissah, Angelina Martin, Raheal Osman, Gloria Dzata and Georgina Banahene, all staff of the National Cardiothoracic Centre at the time of the study, are acknowledged for their assistance with the data collection process.

## Data availability

All relevant data are within the manuscript and its supporting information files

## Supporting information

**S1 Table** Comparison of chronotropic index with stress, anxiety and depressive symptoms by tertiles using the Kruskal-Wallis test

**S2 Table** Comparison of heart rate recovery at 1 minute with stress, anxiety and depressive symptoms by tertiles using one-way ANOVA

**S3 Table** Linear regression model (crude) assessing the association between psychosocial factors and cardiovascular autonomic function

**S4 Table** Quantile regression model (adjusted) assessing the association between psychosocial factors and chronotropic index

**S5 Table** Linear regression model (adjusted) assessing the association between psychosocial factors and HRR_1_

**S6 Table** Linear regression model (adjusted) assessing the association between psychosocial factors and HRR_2_

**S7 Table** Logistic regression model (crude) assessing the association between psychosocial factors and cardiovascular autonomic function

**S8 Table** Logistic regression model (adjusted) assessing the association between psychosocial factors and chronotropic index

**S9 Table** Logistic regression model (adjusted) assessing the association between psychosocial factors and HRR_1_

**S10 Table** Logistic regression model (adjusted) assessing the association between psychosocial factors and HRR_2_

**S11 Table** Study dataset

