## Supplementary material for "Relationship between psychosocial factors and cardiovascular autonomic function among junior doctors: A cross-sectional study": S1 Table

| **PARAMETER** | **TERTILE** | **OBSERVATIONS** | **RANK SUM** | **χ^2^** | **p-value** |
| --- | --- | --- | --- | --- | --- |
| **Stress** | Lowest | 93 | 12320.50 | 3.89 | 0.14 |
|  | Middle | 84 | 9390.00 |  |  |
|  | Highest | 67 | 8179.50 |  |  |
| **Anxiety** | Lowest | 102 | 13051.00 | 1.12 | 0.57 |
|  | Middle | 74 | 8835.50 |  |  |
|  | Highest | 68 | 7997.50 |  |  |
| **Depression** | Lowest | 106 | 13044.00 | 0.03 | 0.99 |
|  | Middle | 66 | 8106.00 |  |  |
|  | Highest | 72 | 8740.00 |  |  |
