## Supplementary material for "Relationship between psychosocial factors and cardiovascular autonomic function among junior doctors: A cross-sectional study": S2 Table

| **Parameter** | **Source** | **Sum of Squares** | **Degrees of freedom** | **Mean square** | **F statistic** | **Significance** |
| --- | --- | --- | --- | --- | --- | --- |
| Stress | Between groups | 336.88 | 2 | 168.44 | 1.90 | 0.15 |
|  | Within groups | 21341.95 | 241 | 88.56 |  |  |
|  | Total | 21678.83 | 243 | 89.21 |  |  |
| Anxiety | Between groups | 67.22 | 2 | 33.61 | 0.37 | 0.69 |
|  | Within groups | 21611.61 | 241 | 89.67 |  |  |
|  | Total | 21678.83 | 243 | 89.21 |  |  |
| Depression | Between groups | 8.91 | 2 | 4.45 | 0.05 | 0.95 |
|  | Within groups | 21669.92 | 241 | 89.91 |  |  |
|  | Total | 21678.83 | 243 | 89.21 |  |  |
