## Supplementary material for "Relationship between psychosocial factors and cardiovascular autonomic function among junior doctors: A cross-sectional study": S3 Table

| Independent variable | R squared | Coefficient | Robust standard error | t | p value | 95% confidence interval |
| --- | --- | --- | --- | --- | --- | --- |
| **Chronotropic index** | | | | | | |
| Stress score | 0.002 | -0.18 | 0.01 | -2.13 | 0.03 | -0.03 – -0.001 |
| Anxiety score | 0.004 | -0.01 | 0.01 | -1.30 | 0.19 | -0.03 – 0.01 |
| Depression score | 0.001 | -0.003 | 0.01 | -0.50 | 0.62 | -0.02 – 0.01 |
| **HRR_1_** | | | | | | |
| Stress score | 0.011 | 1.01 | 0.60 | 1.68 | 0.10 | -1.18 – 2.20 |
| Anxiety score | 0.002 | 0.40 | 0.59 | 0.69 | 0.49 | -0.75 – 1.57 |
| Depression score | <0.001 | 0.14 | 0.53 | 0.27 | 0.79 | -0.90 – 1.18 |
| **HRR_2_** | | | | | | |
| Stress score | 0.008 | 0.87 | 0.60 | 1.45 | 0.15 | -0.31 – 2.05 |
| Anxiety score | <0.001 | 0.11 | 0.63 | 0.18 | 0.86 | -1.12 – 1.34 |
| Depression score | 0.008 | 0.86 | 0.59 | 1.46 | 0.15 | -0.30 – 2.02 |
