## Supplementary material for "Relationship between psychosocial factors and cardiovascular autonomic function among junior doctors: A cross-sectional study": S4 Table

Observations – 244

Raw sum of deviations – 8.96

Minimum sum of deviations – 7.85

Pseudo R^2^ – 0.12

| Independent variable | Coefficient | Robust standard error | t | p value | 95% confidence interval |
| --- | --- | --- | --- | --- | --- |
| Stress score | -0.10 | 0.01 | -1.38 | 0.17 | -0.03 – 0.00 |
| Anxiety score | -0.00 | 0.01 | -0.41 | 0.68 | -0.02 – 0.01 |
| Depression score | 0.01 | 0.01 | 0.07 | 0.50 | -0.01 – 0.02 |
| Age | 0.00 | 0.00 | 1.14 | 0.26 | -0.001 – 0.004 |
| Sex (male) | -0.71 | 0.01 | -5.15 | <0.01 | -1.00 – -0.04 |
| BMI | 0.00 | 0.01 | 0.83 | 0.41 | -0.001 – 0.003 |
| Smoking | 0.30 | 0.06 | 0.47 | 0.64 | -0.09 – 0.14 |
| Coffee intake | 0.00 | 0.01 | 0.01 | 1.00 | -0.03 – 0.03 |
| Resting heart rate | 0.00 | 0.00 | 2.32 | 0.02 | 0.0001 – 0.0023 |
