## Supplementary material for "Relationship between psychosocial factors and cardiovascular autonomic function among junior doctors: A cross-sectional study": S5 Table

Observations – 244

R(9,234) – 2.11

R^2^ – 0.11

Root mean square error – 9.09

p-value – 0.03

| Independent variable | Coefficient | Robust standard error | t | p value | 95% confidence interval |
| --- | --- | --- | --- | --- | --- |
| Stress score | 1.53 | 0.76 | 2.01 | 0.045 | 0.03 – 3.02 |
| Anxiety score | -0.08 | 0.70 | -0.11 | 0.915 | -1.46 – 1.31 |
| Depression score | -0.60 | 0.62 | -0.98 | 0.329 | -1.82 – 0.61 |
| Age | -0.13 | 0.13 | -0.98 | 0.330 | -0.39 – 0.13 |
| Sex (male) | -2.61 | 1.27 | -2.05 | 0.041 | -5.12 – -0.11 |
| BMI | -0.06 | 0.13 | -0.42 | 0.626 | -0.32 – 0.21 |
| Smoking | -3.62 | 4.04 | -0.89 | 0.372 | -11.57 – 4.34 |
| Coffee intake | -0.54 | 1.20 | -0.45 | 0.653 | -2.91 – 1.82 |
| Resting heart rate | -0.23 | 0.07 | -3.33 | 0.001 | -0.37 – -0.09 |
