## Supplementary material for "Relationship between psychosocial factors and cardiovascular autonomic function among junior doctors: A cross-sectional study": S6 Table

Observations – 244

R (9,234) – 3.23

R^2^ – 0.14

Root mean square error – 9.39

p-value – <0.01

| Independent variable | Coefficient | Robust standard error | t | p value | 95% confidence interval |
| --- | --- | --- | --- | --- | --- |
| Stress score | 0.82 | 0.84 | 0.98 | 0.33 | -0.83 – 2.47 |
| Anxiety score | -0.91 | 0.80 | -1.13 | 0.26 | -2.49 – 0.67 |
| Depression score | 0.93 | 0.75 | 1.24 | 0.22 | -0.55 – 2.41 |
| Age | -0.28 | 0.13 | -2.13 | 0.03 | -0.53 – -0.02 |
| Sex (male) | 2.18 | 1.32 | -1.65 | 0.10 | -4.18 – 0.43 |
| BMI | -0.04 | 0.14 | -0.25 | 0.80 | -0.32 – 0.25 |
| Smoking | 3.48 | 3.30 | 1.05 | 0.29 | -3.02 – 9.97 |
| Coffee intake | -0.94 | 1.24 | -0.76 | 0.45 | -3.40 – 1.51 |
| Resting heart rate | -0.27 | 0.07 | -3.67 | <0.01 | -0.41 – -0.12 |
