## Supplementary material for "Relationship between psychosocial factors and cardiovascular autonomic function among junior doctors: A cross-sectional study": S7 Table

| Independent variable | Pseudo R squared | Odds ratio | Robust standard error | z | p value | 95% confidence interval |
| --- | --- | --- | --- | --- | --- | --- |
| **Chronotropic index** | | | | | | |
| Stress score | 0.001 | 1.21 | 0.26 | 0.92 | 0.36 | 0.80 – 1.86 |
| Anxiety score | 0.003 | 1.15 | 0.24 | 0.66 | 0.51 | 0.76 – 1.74 |
| Depression score | 0.010 | 0.74 | 0.20 | -1.14 | 0.25 | 0.44 – 1.25 |
| **HRR_1_** | | | | | | |
| Stress score | 0.019 | 0.65 | 0.25 | 1.11 | 0.27 | 0.30 – 1.40 |
| Anxiety score | <0.001 | 1.06 | 0.37 | 0.17 | 0.87 | 0.53 – 2.12 |
| Depression score | 0.038 | 0.42 | 0.27 | -1.34 | 0.18 | 1.12 – 1.49 |
| **HRR_2_** | | | | | | |
| Stress score | 0.002 | 0.89 | 0.24 | -0.44 | 0.66 | 0.52 – 1.51 |
| Anxiety score | 0.001 | 1.09 | 0.28 | 0.33 | 0.74 | 0.66 – 1.82 |
| Depression score | 0.025 | 0.56 | 0.22 | -1.46 | 0.14 | 0.26 – 1.22 |
