## Supplementary material for "Relationship between psychosocial factors and cardiovascular autonomic function among junior doctors: A cross-sectional study": S8 Table

Observations – 244

Pseudo R^2^ – 0.09

p-value – 0.12

| Independent variable | Adjusted odds ratio | Standard error | z | p value | 95% confidence interval |
| --- | --- | --- | --- | --- | --- |
| Stress score | 1.68 | 0.51 | 1.73 | 0.08 | 0.93 – 3.04 |
| Anxiety score | 1.30 | 0.38 | 0.88 | 0.38 | 0.73 – 2.31 |
| Depression score | 0.44 | 0.16 | -2.30 | 0.02 | 0.22 – 0.88 |
| Age | 0.91 | 0.05 | -1.59 | 0.11 | 0.82 – 1.02 |
| Sex (male) | 2.47 | 1.27 | 1.76 | 0.08 | 0.90 – 6.77 |
| BMI | 1.00 | 0.05 | 0.03 | 0.98 | 0.91 – 1.10 |
| Smoking | 1.09 | 1.36 | 0.07 | 0.95 | 0.09 – 12.63 |
| Coffee intake | 0.89 | 0.42 | -0.25 | 0.80 | 0.35 – 2.24 |
| Resting heart rate | 1.04 | 0.02 | 1.74 | 0.08 | 1.0 – 1.08 |
