## Supplementary material for "Relationship between psychosocial factors and cardiovascular autonomic function among junior doctors: A cross-sectional study": S9 Table

Observations – 238 (smoking omitted due to multiple collinearity)

Pseudo R^2^ – 0.34

p-value – 0.002

| Independent variable | Adjusted odds ratio | Standard error | z | p value | 95% confidence interval |
| --- | --- | --- | --- | --- | --- |
| Stress score | 0.68 | 0.49 | -0.53 | 0.59 | 0.17 – 2.79 |
| Anxiety score | 2.29 | 1.48 | 1.28 | 0.20 | 0.64 – 8.11 |
| Depression score | 0.11 | 0.15 | -1.66 | 0.10 | 0.01 – 1.50 |
| Age | 1.02 | 0.11 | 0.17 | 0.86 | 0.83 – 1.25 |
| Sex (male) | 3.14 | 3.08 | 1.17 | 0.24 | 0.46 – 21.47 |
| BMI | 0.97 | 0.09 | -0.37 | 0.71 | 0.80 – 1.16 |
| Coffee intake | 1.36 | 0.11 | 0.35 | 0.72 | 0.25 – 7.32 |
| Resting heart rate | 1.18 | 0.06 | 3.20 | <0.01 | 1.07 – 1.31 |
