## Supplementary material for "Relationship between psychosocial factors and cardiovascular autonomic function among junior doctors: A cross-sectional study": S10 Table

Observations – 238 (smoking omitted due to multiple collinearity)

Pseudo R^2^ – 0.12

p-value – 0.10

| Independent variable | Adjusted odds ratio | Standard error | z | p value | 95% confidence interval |
| --- | --- | --- | --- | --- | --- |
| Stress score | 1.13 | 0.45 | 0.31 | 0.757 | 0.52 – 2.48 |
| Anxiety score | 1.62 | 0.61 | 1.29 | 0.191 | 0.78 – 3.39 |
| Depression score | 0.34 | 1.19 | -1.97 | 0.049 | 0.11 – 0.997 |
| Age | 0.99 | 0.07 | -0.18 | 0.856 | 0.87 – 1.12 |
| Sex (male) | 1.72 | 1.06 | 0.88 | 0.380 | 0.51 – 5.77 |
| BMI | 0.95 | 0.06 | -0.81 | 0.418 | 0.84 – 1.08 |
| Coffee intake | 1.38 | 0.78 | 0.57 | 0.570 | 0.45 – 4.19 |
| Resting heart rate | 1.07 | 0.03 | 2.54 | 0.011 | 1.02 – 1.12 |
